# Ambulatory detection of isolated REM sleep behavior disorder combining actigraphy and questionnaire

**DOI:** 10.1101/2022.04.21.22274011

**Authors:** Andreas Brink-Kjaer, Niraj Gupta, Eric Marin, Jennifer Zitser, Oliver Sum-Ping, Anahid Hekmat, Flavia Bueno, Ana Cahuas, James Langston, Poul Jennum, Helge B.D. Sorensen, Emmanuel Mignot, Emmanuel During

## Abstract

**Background:** Isolated rapid-eye-movement sleep behavior disorder (iRBD) is in most cases a prodrome of neurodegenerative synucleinopathies, affecting 1-2% of middle-aged and older adults, however accurate ambulatory diagnostic methods are lacking. Questionnaires lack specificity in non-clinic populations. Wrist actigraphy can detect characteristic features in individuals with RBD, however high frequency actigraphy has rarely been used.

**Objectives:** To develop a machine learning classifier using high frequency (1-second resolution) actigraphy and a short patient survey for detecting iRBD with high accuracy and precision.

**Methods:** Analysis of ≥7 nights home actigraphy data and 9-item questionnaire (RBD Innsbruck inventory and 3 synucleinopathy prodromes of subjective hyposmia, constipation and orthostatic dizziness) in a dataset including 42 patients with iRBD, 21 sleep clinic patients with other sleep disorders, and 21 community controls.

**Results:** The actigraphy classifier achieved 95.2% (95% CI: 88.3 - 98.7) sensitivity and 90.9% (95% CI: 82.1 - 95.8) precision. The questionnaire classifier achieved 90.6% accuracy and 92.7% precision, exceeding performance of RBD-I and prodromal questionnaire alone. Concordant predictions between actigraphy and questionnaire reached specificity and precision of 100% (95% CI: 95.7 - 100.0) with 88.1% sensitivity (95% CI: 79.2 - 94.1) and outperformed any combination of actigraphy plus single question on RBD or prodromal symptoms.

**Conclusions:** Actigraphy detected iRBD with high accuracy in a mixed clinical and community cohort. This cost-effective fully remote procedure can be used to diagnose iRBD in specialty outpatient settings and has potential for large scale screening of iRBD in the general population.

## 1 Introduction

Rapid-eye-movement (REM) sleep behavior disorder (RBD) is a motor disinhibition during REM sleep that causes frequent twitches, jerks, and unpredictable, often violent episodes of dream enactment^1^. In most cases, RBD is associated with an underlying alpha-synucleinopathy, more commonly Parkinson’s disease or Dementia with Lewy bodies^2^. In this setting, RBD and other prodromes, such as autonomic impairment (constipation and orthostasis) and hyposmia, can precede overt neurodegenerative disease by a decade or more^3,4^. Early diagnosis of “isolated RBD” (iRBD, formerly called idiopathic RBD)^5,6^ reduces risk of injuries through implementation of safety measures^7^ and provides lead time for neuroprotective interventions.

In the general population, the prevalence of iRBD is 1-2 % of middle-aged and older adults. In neurology and sleep clinics, the prevalence may be even higher, however diagnosing iRBD clinically is often challenging. Video-polysomnography (vPSG) provides a definitive diagnosis, but is a complex, time- and resource-intensive procedure, mostly available in academic sleep centers^8^. Further, an overnight stay in the sleep lab is not acceptable to all patients, and a single night stay may not be sufficient to capture dream-enactment behaviors. RBD questionnaires are a convenient initial step to diagnose iRBD but focus on the behavioral symptoms, which lack specificity since common conditions, such as obstructive sleep apnea, periodic limb movements, and non-REM parasomnias can present similarly. Although significantly higher than in non-clinical settings^9–12^, the positive predictive value of RBD questionnaires in clinic populations does not exceed 80%, which is arguably insufficient for screening a condition with such serious implications.

Actigraphy can be used to assess nighttime activity. Actigraphs contain accelerometers measuring movement, which have been used for over three decades as a noninvasive method for evaluating sleep and circadian disturbances^13,14^. However, studies using nocturnal actigraphy show inconsistent performance for distinguishing RBD from other conditions^15–17^. Limitations may be related to a reductionist approach comparing a single feature of movements (total activity count per night, activity frequency, etc.) while neglecting other characteristics of RBD, including its’ time distribution. One study described patterns of high activity coinciding with REM sleep and a feature called the “short burst inactivity” index, which was found to be elevated in iRBD^17^. A later study^18^ found that this feature alone could reach > 80 % accuracy by analyzing data from high-frequency actigraphy in 1-second windows. Conceivably, actigraphy could become a convenient ambulatory procedure in clinic populations with a high pretest probability.

In this study, we tested a new diagnostic procedure of iRBD combining the well-established Innsbruck RBD Inventory, a brief 3-item questionnaire screening for common synucleinopathy prodromes (olfactory loss, constipation, and orthostatic symptoms), and ambulatory high frequency actigraphy assisted by machine learning.

## 2 Methods

### 2.1 Participants

Participants were enrolled between April and December 2021. IRBD cases and clinical controls (CL) were recruited in the Stanford Sleep Center, and community CL were recruited via online advertisement. IRBD cases required a definitive diagnosis according to the International Classification of Sleep Disorders, 3^rd^ edition (ICSD-3) criteria^19^, excluding participants with narcolepsy, overt synucleinopathy, or dementia. For the clinical CL set, goal was to include at least 25 % subjects with NREM parasomnias, 25% with untreated moderate or severe OSA, and 25% with restless legs syndrome (RLS) or PLM >15 per hour of sleep. Clinic CL required unequivocal absence of REM sleep without atonia (RSWA) within 3 years of study participation. Community CL were enrolled consecutively without predetermined ratio of sleep pathology, however, to minimize risk of enrolling participants with unknown RBD, they could not have a history of dream enactment. We estimated a sample size of 40 in each group based on a prior study^18^. This study was approved by the Institutional Review Board of Stanford and all participants provided written informed consent.

### 2.2 Procedure

Participants received a study packet containing an AX-6 (Axivity, Ltd) device recording at 25Hz and dynamic range ±8 gravity (g), sleep logs and questionnaires. They were asked to wear the actigraph on their dominant wrist for at least 14 nights, complete sleep diary, parasomnia log reporting any abnormal behaviors during sleep, the Innsbruck RBD inventory (RBD-I) and summary question^20^, and a novel 3-item questionnaire (3Q) on other prodromal synucleinopathy symptoms: known or subjective hyposmia, constipation and orthostasis (see eMethods). Participants could continue their usual treatments and mailed back their study packet upon completion of the procedure.

### 2.3 Actigraphy

Actigraphy data was included in the analysis only if the device recorded at least 7 nights and all questionnaires and forms were fully completed. Data was edited to keep sleep periods based on sleep diaries.

#### 2.3.1 Activity Count

The activity count was calculated in 1-second, 15-second, and 30-second bins according to the procedure shown in eMethods.

#### 2.3.2 Actigraph Features

A set of features were derived to best capture activity that distinguish iRBD cases from CL. These include:

1. Features derived from the sleep diary: Total sleep time (TST); wake after sleep onset (WASO); sleep efficiency (SE), TST / (TST + WASO); sleep onset; and sleep offset.
2. Features from prior studies^17,18^, with varying parameters enabling a more data-driven search for optimal accuracy: Activity index (AI), % x-second epochs with any activity for x ∈ {10, 30, 60}; short immobile bouts (SIB), (# zero-activity periods larger than x seconds and less than 60 seconds) / TST for x ∈ {0, 1, 5}; and mean motor activity score (MMAS), average activity count.
3. Novel features that capture new RBD activity patterns, based on visual interpretation of the data from the first 15 consecutive cases and 5 clinic CL: Twitch activity (TA), (# activity counts with larger than 0 and smaller than x and with surrounding zero-activity) / TST for x ∈ {0.5, 1, 1.5}; long immobile bouts (LIB), (# zero-activity periods longer than x seconds) / TST for x ∈ {60, 120, 300}; Hjorth parameters of activity counts [Hjorth activity (HPA), Hjorth mobility (HPM), and Hjorth Complexity (HPC)]^21^.

#### 2.3.3 Feature windows

Loss of skeletal muscle atonia and propensity for sudden motor activity during REM sleep would be expressed as periods of higher activity counts on actigraphy. However, frequent movements can also occur in NREM sleep due to PLM and OSA, and during brief periods of wake as sleep drive gradually reduces. Actigraphy cannot distinguish between sleep stages. To minimize the risk of analyzing NREM and wake periods, and maximize likelihood of including one to two REM cycles, we applied an empiric time window labelled “possible REM sleep” (*pREM*) starting 70 minutes, and ending 200 minutes after reported sleep onset. We also labeled periods as “wake” based on sleep diaries. We computed features based on data within these windows by: 1) excluding or keeping wake windows; 2) excluding, keeping, or only keeping *pREM*. In total, this corresponds to 6 feature variations. Fig. 1 shows examples of actigraphy data together win the pREM window and hypnograms derived from concurrent PSG.

**Figure 1.**
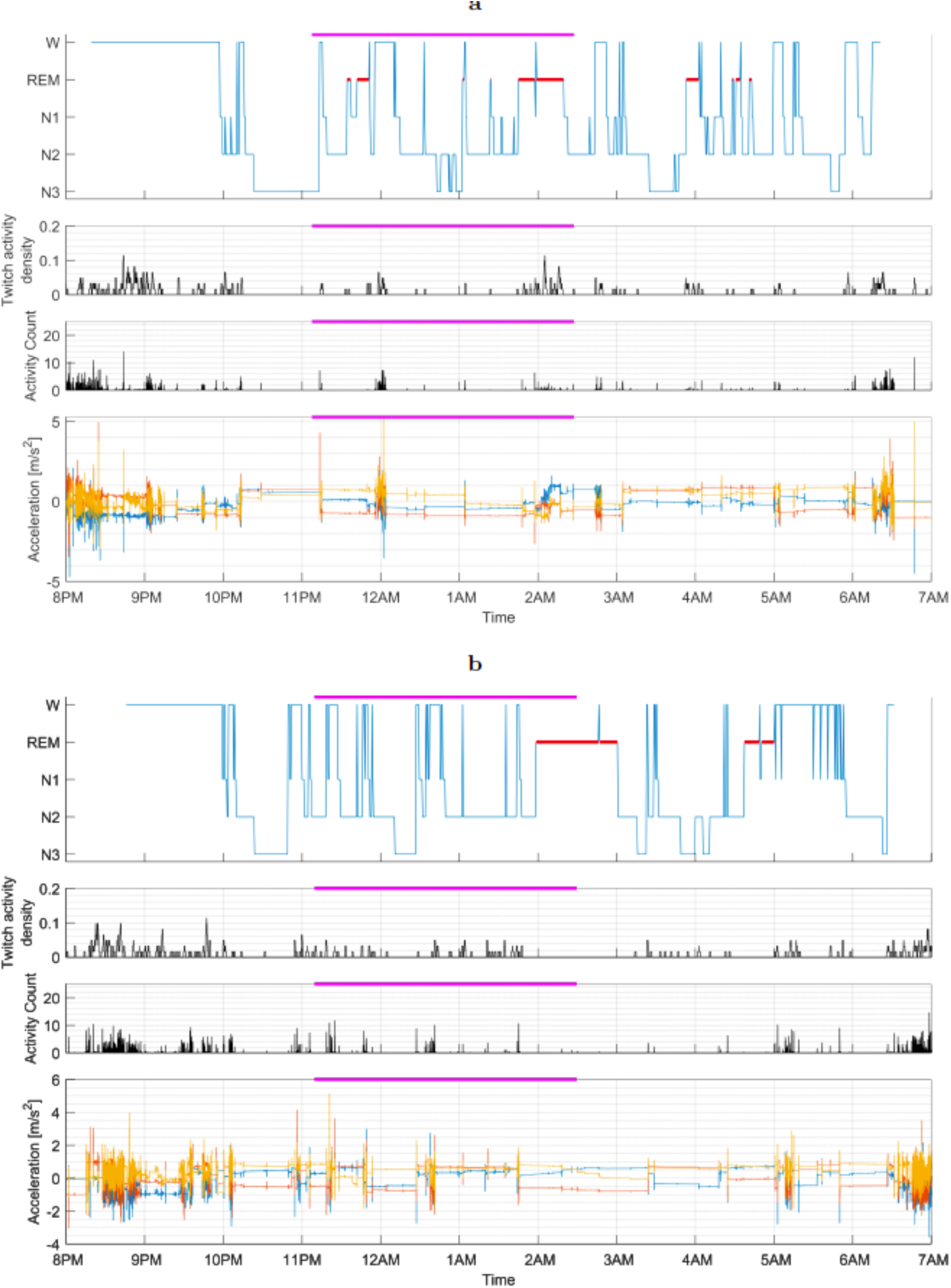
Concurrent actigrams and hypnograms recorded during an in-lab video-polysomnography in participants with and without isolated RBD. The data displayed are from (a) one iRBD case and (b) one control. The magenta line marks the “probable REM sleep” window (70-200 minutes after estimated sleep onset). The density of twitch activity is visualized by applying a moving average filter of 61 seconds to events of isolated activity counts less than 0.5 in amplitude.

### 2.4 Classification Models

We developed a framework for classification of iRBD vs. non-iRBD based on a single night of actigraphy which outputs an iRBD prediction score (“per night” prediction). Subsequently, all output iRBD scores for a same subject can be averaged, resulting in a single prediction (“per subject” prediction). Finally, actigraphy prediction could be compared to questionnaire-based prediction.

We compared the traditional single-feature approach against a large set of features in a machine learning model (multi-feature model). Specifically, we fitted a threshold for each feature and an ensemble of decision trees using all derived features (see eMethods). For comparison with prior methods, the single feature selected would be the SIB^17,18^, fitted to our data based on the lowest *p*-value in group comparisons using Mann-Whitney tests.

Similarly, this approach was applied to questionnaire data, which contained nine items: the Innsbruck summary question, five RBD-I items, and three 3Q items for subjective hyposmia, constipation, and orthostasis. Machine learning models were optimized for the 3Q only, the Innsbruck RBD-I and summary question only, and all questions together. In case of “don’t know” answers, a value of 0.5 was imputed for the boosted decision trees. The decision trees are invariant to the exact imputation value if it is between 0 and 1.

#### 2.4.1 Validation

The classifiers were fitted and evaluated in a leave-one-out cross validation setup. Iteratively, all nights of actigraphy from a participant were held out while fitting, subsequently, the model was validated in the held-out data. Hence, each model was fitted as many times as the number of participants, resulting in an unbiased performance estimate in all subjects.

Once both actigraphy and questionnaire models were developed and validated, a two-dimensional model was created. This 3-class classifier would output iRBD, non-iRBD, or “undetermined diagnosis”, in case of concordant positive, concordant negative, or discordant prediction, respectively. The performance of actigraphy combined with single item questions, i.e., RBD-I summary question, hyposmia, constipation, and orthostasis would also be evaluated.

### 2.5 Statistical Tests

Statistical analyses were performed in Matlab (R2019b, The Mathworks). Group comparisons were carried out using chi-squared test for nominal variables and Mann-Whitney tests for numerical variables. Classifiers were evaluated using sensitivity, specificity, precision, and accuracy. Additionally, receiver-operating-curves (ROC) analyses were performed to calculate the area under the ROC curve (AUC). Performance confidence intervals were estimated using Clopper-Pearson’s method.

Correlations were evaluated between individual actigraphy features as well as overall actigraphy prediction score and REM sleep without atonia index (RSWAi), periodic limb movements index (PLMi), apnea-hypopnea index (AHI), sleep diagnoses, and RBD treatments.

Performance was evaluated as a function of number of nights recorded with actigraphy. Due to the wide range of nights recorded, we restricted this analysis to recordings with at least 14 nights. Thereby, the sample we consider in this analysis do not change depending on the number of nights analyzed. Confidence bounds were estimated using bootstrapping in 1000 iterations, i.e., for each participant, the performance was evaluated in subsets of nights that were randomly sampled.

## 3 Results

Out of the initial 110 participants initially selected, 26 were excluded: 9 due to an uncertain diagnosis, including borderline RSWAi (n=4), history of clinical RBD but absence of a PSG report documenting abnormal RSWA (n=3), secondary report of dream-enactment behaviors in community CL participants (n=2), and 16 due to noncompliance with study procedure (n=11), device failure or loss of device (n=5). Additionally, one participant with iRBD was excluded after she reported receiving sodium oxybate treatment, a REM suppressant drug. The final dataset used for the analysis comprised 84 participants: 42 iRBD cases and 42 CL, including 21 clinic and 21 community CL (Table 1).

**Table 1:** Summary of demographic and clinical data. Statistics are computed for entire control groups combined using chi-squared tests for sex, antidepressants, RLS, NREM parasomnia, hypersomnia, and OSA, while Mann-Whitney tests for other variables. In case of missing data, the sample size is denoted below the variable. *P*-values < 0.05 are considered significant and marked in bold. PLMi: periodic leg movement index; AHI: apnea-hypopnea index; NREM: non-rapid eye movement; OSA: obstructive sleep apnea; PAP: positive airway pressure therapy; RLS/PLM: restless legs syndrome/periodic limb movements, as defined by PLMi≥15; RSWAi: REM sleep without atonia index according to ICSD-3 criteria^19^.

|  |  | iRBD | Clinic Controls | Community controls | *p*-value |
| --- | --- | --- | --- | --- | --- |
|  | n | 42 | 21 | 21 |  |
| Age | $\mu \pm \sigma$ | 67.3 ± 6.69 | 63.1 ± 11.8 | 65.5 ± 8.6 | 0.28 |
| BMI | $\mu \pm \sigma$ | 26.3 ± 3.57 | 28.6 ± 5.3 | 23.4 ± 3.3 | 0.792 |
| Sex (% male) | n, (%) | 33, (78.6 %) | 16, (76.2 %) | 13, (61.9 %) | 0.321 |
| Antidepressants | n, (%) | 12, (28.6 %) | 2, (9.5 %) | 2, (4.8 %) | **0.00341** |
| Melatonin | n, (%) | 29, (69.0 %) | 0, (0 %) | 0, (0 %) | **2.83$\times 10^{-11}$** |
| Clonazepam | n, (%) | 18, (42.9 %) | 1, (4.8 %) | 0, (0 %) | **9.27$\times 10^{-6}$** |
| Rivastigmine | n, (%) | 6, (14.3 %) | 0, (0 %) | 0, (0 %) | **0.011** |
| Pramipexole | n, (%) | 3, (7.14 %) | 0, (0 %) | 0, (0 %) | 0.0778 |
| PLMi | $\mu \pm \sigma$ | 30.9 ± 38.3 | 12.1 ± 17.0<br>(n=18) | - | **0.0224** |
| OSA on PAP therapy | n, (%) | 11, (26.2 %) | 8, (38.1 %) | 0, (0 %) | 0.434 |
| AHI in untreated OSA | $\mu \pm \sigma$ | 10.8 ± 8.65<br>(n=31) | 15.2 ± 10.5<br>(n=14) | 9.0 ± 3.6<br>(n=3) | 0.27 |
| RLS/PLM | n, (%) | 3, (7.14 %) | 6, (28.6 %) | 1, (4.8 %) | 1 |
| NREM parasomnia | n, (%) | 1, (2.38 %) | 5, (23.8 %) | 0, (0 %) | 0.0901 |
| Hypersomnia | n, (%) | 6, (14.3 %) | 2, (9.5 %) | 1, (4.8 %) | 0.29 |
| Insomnia | n, (%) | 2, (4.76 %) | 3, (14.3 %) | 4, (19.0 %) | 0.0778 |
| Parasomnia episodes per night | $\mu \pm \sigma$ | 0.921 ± 1.28 | 0.03 ± 0.09<br>(n=5) | 0 ± 0 | **1.5$\times 10^{-12}$** |
| RSWai | $\mu \pm \sigma$ | 0.66 ± 0.26<br>(n=36) | - | - | - |

No between-group differences were observed for age, sex, and BMI. Clinic CL included 5 with NREM parasomnias, 7 untreated OSA (4 moderate, 3 severe) and 6 with RLS/PLM. Cases and CL sets were similar in terms of usage of PAP device and AHI. As expected, iRBD cases had higher PLMi and were more likely to use melatonin and clonazepam than CL. Other RBD treatments included transdermal rivastigmine and pramipexole. Antidepressant use was more frequent in iRBD.

On average, 17.6 ± 8.1 and 25.2 ± 8.2 nights (SD) were recorded in the CL and iRBD groups, respectively. Due to the heterogeneity within each group, the classification performances were evaluated as a function of number of nights.

### 3.1 Actigraphy features and questionnaire

Two features were found to have higher *p*-values than prior feature SIB: LIB, TA (see eTable 1 for details).

All questions of the RBD-I and 3Q showed significance (*p*-value < 0.05). Among all questions, the most accurate was RBD-I item 3 (sleep-related extensive movements) with 93.6 % accuracy (eTable 2).

### 3.2 iRBD Classification

The multi-feature actigraphy classifier achieved best performance using 1-second windows, with 92.9 % accuracy and 90.9% precision (AUC 0.977) analyzing all nights (see Fig. 2 and Table 2).

**Figure 2.**
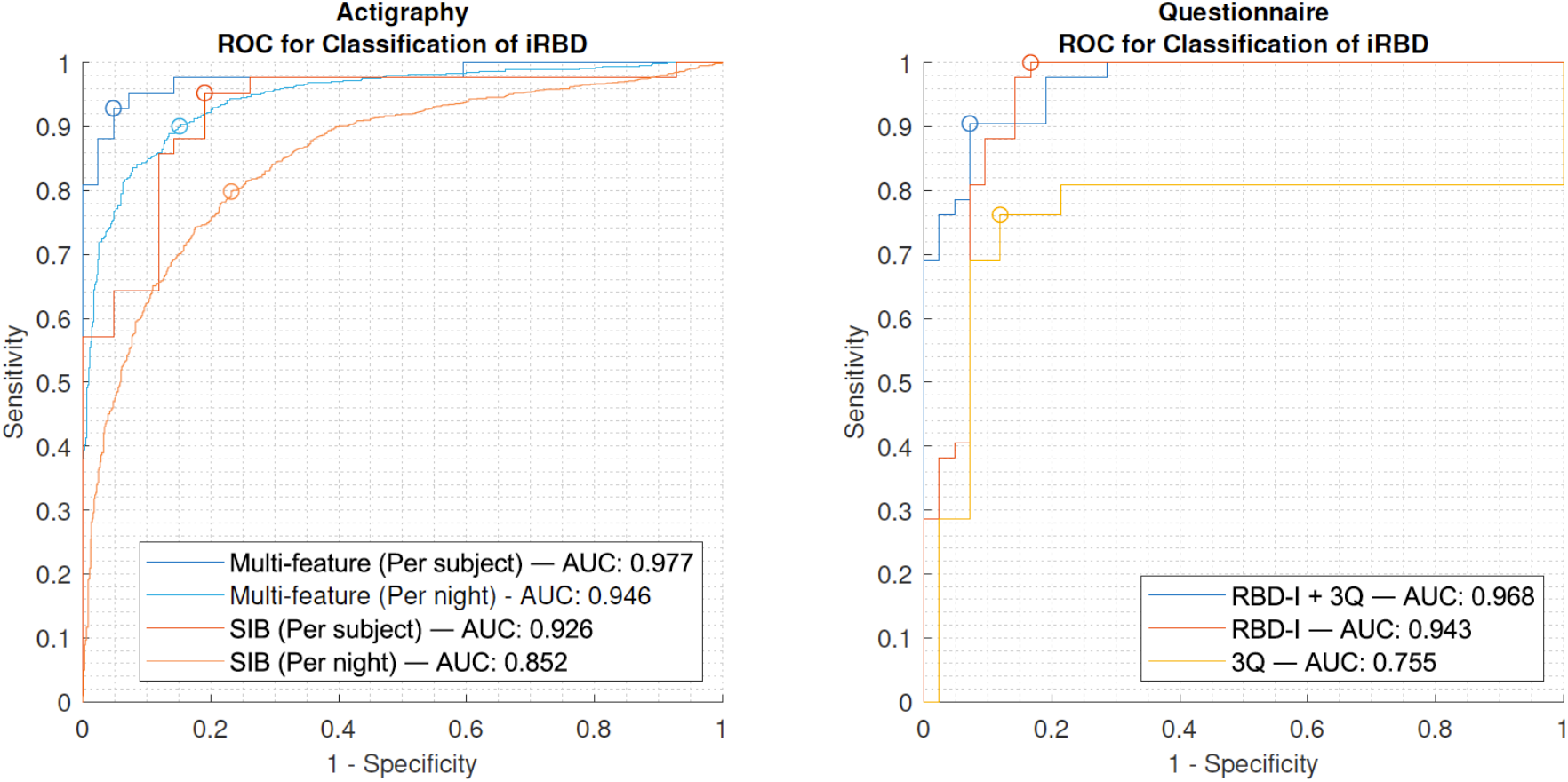
ROC for iRBD classification using actigraphy or questionnaires. The ROC curves for classification using actigraphy are generated per subject and per night for the boosted decision trees (multi-feature) and SIB (single-feature). ROC: receiver-operator-characteristics; iRBD: isolated REM sleep behavior disorder; SIB: short immobile bouts; AUC: area under the ROC curve; RBD-I: Innsbruck RBD inventory; 3Q: 3 prodromal symptom questions.

**Table 2:**
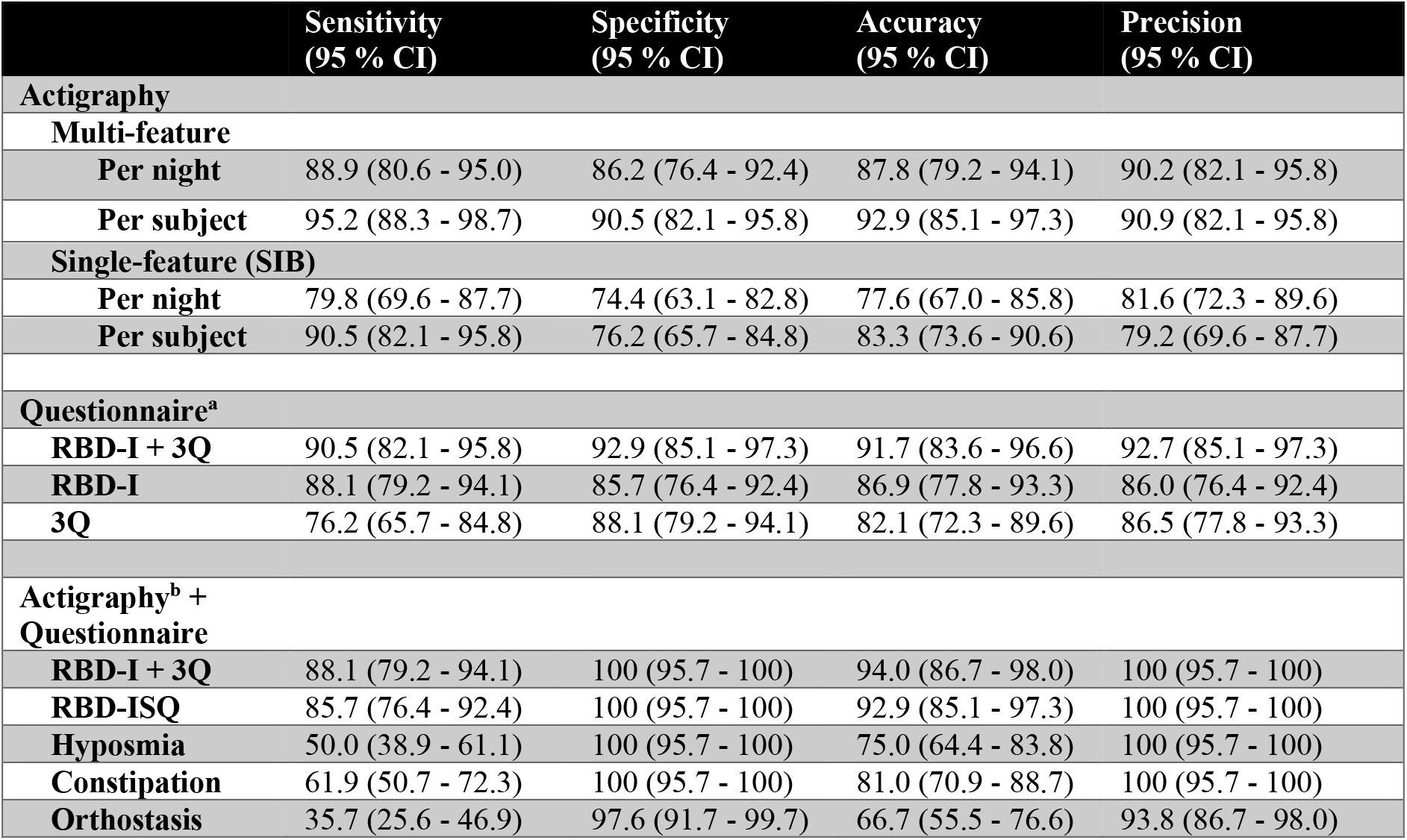
Performance in classification of isolated REM sleep behavior disorder. The performance is reported for classification based on actigraphy, questionnaires, and the combination of these. The combination classifies a sample as belonging to the iRBD group if both classifiers are positive. The 95 % CI were estimated using the Clopper-Pearson method. CI: confidence interval; SIB: short immobile bouts; RBD-I: Innsbruck RBD inventory (5 items + summary question) ; RBD-ISQ: Innsbruck RBD inventory summary question; 3Q: 3-item prodromal synucleinopathy questionnaire (hyposmia, constipation, orthostasis); a: machine learning classifier; b: actigraphy analysis using machine learning multi-feature model and all recorded nights (“per subject”);

Optimizing threshold (eTable 3) improved accuracy to 94.0 % using all nights. Analysis in 15- or 30-second windows both yielded similar accuracies of 91.7% and 92.9%, respectively (eTable 4 and 5).

In comparison, the single-feature SIB classifier achieved 83. 3% accuracy and 79.2 % precision (AUC 0.926) with all nights, after optimization using the parameter (x = 5) and a median cut-off 0.379 across cross-validation folds.

The machine learning classifier using all questionnaire data (6-item RBD-I + prodromal 3Q) performed with 91.7 % accuracy and 92.7 % precision (AUC 0.968). Restricting the analysis to either the RBD-I or 3Q achieved 86.9 % and 82.1 % accuracy, respectively.

The two-dimensional model combining actigraphy and questionnaires achieved 88.1 % sensitivity and 100 % precision (see Table 2 and Fig. 3). Predictions were concordant in 90.5 % of cases and 83.3 % of CL. In 9.5 % (*n* = 4) of cases, the output diagnosis was undetermined, and 2.3 % (*n* = 1) of cases were misdiagnosed as non-iRBD. In 16.7 % (*n* = 7) of CL, the output diagnosis was undetermined.

**Figure 3.**
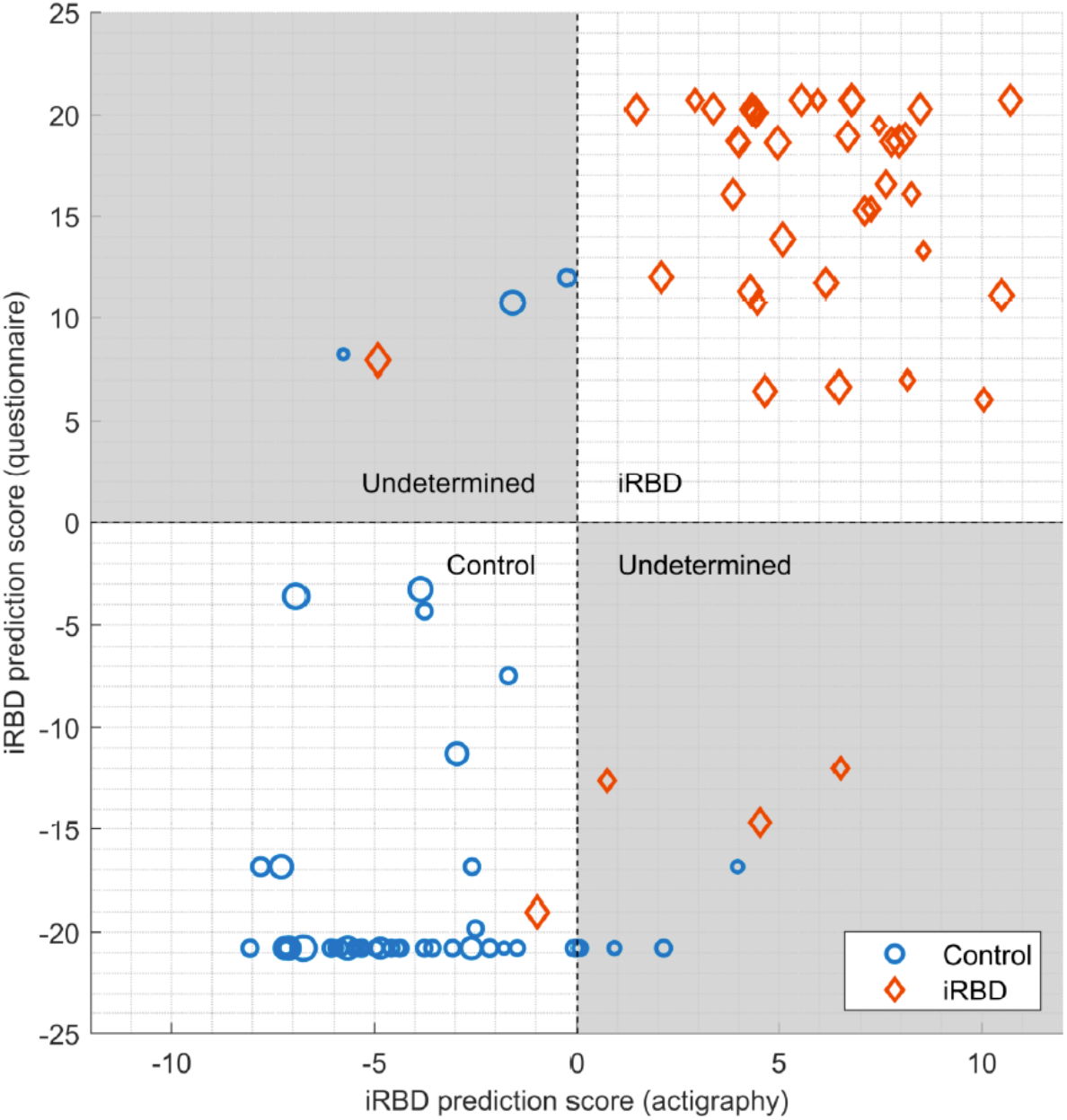
Classifier iRBD scores based on actigraphy and Innsbruck RBD questionnaire. The dashed lines indicate the default decision lines for each classifier and the gray area signifies “undetermined diagnosis”. The size of each marker is scaled to the number of nights recorded with actigraphy. The iRBD prediction score from questionnaire used the Innsbruck RBD-I and 3 prodromal symptom questions.

No CL was misdiagnosed as RBD. Combination of actigraphy and single item questions yielded the following sensitivity and specificity: 85.7% and 100% with RBD-I summary question, 50% and 100% with subjective hyposmia, 57.1% and 100% with constipation, and 35.7% and 97.6% with orthostasis.

Analysis of incremental performance over multiple nights showed gain in sensitivity and specificity over 7 and 10 nights, respectively, after which performance only marginally improved (eFig. 1).

#### 3.3 Interpretation of models

In the CL group, hypersomnia and insomnia correlated with higher RBD prediction score (hypersomnia: *r* = 0.36, *p* = 0.018; insomnia: *r* = 0.36, *p* = 0.019). No correlation was found between RSWAi or number of RBD episodes, and prediction score.

TA and LIB were the two features with highest importance in the multi-feature actigraphy model (eFig. 2). None of the feature masks had more importance than the whole-night window.

In the questionnaire model, the RBD-I item 1 (violent/aggressive dream content), showed the highest importance (eFig. 3).

## 4 Discussion

IRBD is rarely diagnosed before injuries to self or bed partners occur or the disorder converts to a defined synucleinopathy. Even for sleep experts, diagnosing RBD based on history alone can be challenging, and vPSG is not easily accessible or accepted by patients. In this study, we developed a multimodal iRBD detection framework combining a 9-item questionnaire screening for signs of RBD and prodromal symptoms of synucleinopathy with ambulatory actigraphy, in a dataset of cases and CL from the sleep clinic and community.

Using an ensemble of decision trees, actigraphy detected iRBD with 95.2% sensitivity and 90.5% specificity. Concordant iRBD prediction between actigraphy and questionnaire models had 88.1% sensitivity and 100% precision with performance plateauing out after 7 to 10 nights. Analysis in 1-, 15- and 30-seconds windows did not affect the performance much resulting in accuracies of 92.9%, 91.7 %, and 92.9%.

We found that single-feature actigraphy using SIB had lower accuracy close to 83 %, consistent with prior report^18^. We also designed two actigraphy features with higher discriminative value, LIB and TA. Video-based studies show that in RBD a majority of movements are simple, non-purposeful twitches more prominent in the arms^22–25^. We found that the periodicity of movements detected at the wrist is variable, unlike in other pathologies.

No relation was found between RSWAi or clinical RBD activity, and iRBD prediction scores. This could be due to the small sample size, or to a more complex relation between RSWA, covert (small twitches), and overt (clinical episodes) manifestations in RBD. If the detection model truly operates independent of the RSWA burden and clinical severity, it could potentially identify individuals with borderline RSWA, or with RSWA but no overt manifestation. This entity coined “prodromal RBD” is of great importance as it carries a high lifetime risk of synucleinopathy but is rarely detected^26^. The 4 CL misclassified by actigraphy included one with insomnia and OSA on PAP therapy, one with untreated REM-related OSA (AHI = 19), history of observed leg kicking in sleep and PLMi of 8, and two community CL without vPSG data – one with occasional sleep talking and one with untreated mild-moderate OSA (AHI = 12) and hypersomnia. It shows that actigraphy has possible RBD mimics that would be difficult to exclude with this modality alone.

The comprehensive 9-item questionnaire model achieved 91.7 % accuracy, outperforming models using only the RBD-I or 3Q prodromal questionnaire (86.9 % and 82.1 % accuracy, respectively). Considered alone, the performance of the RBD-I classifier was not superior to that of the traditional cutoff method^17,20^. The prevalence of synucleinopathy prodromes in the CL group was comparable to other studies, with 21% endorsing at least one prodrome and 7%, two or more^27,28^. Among iRBD patients, 52.4% reported subjective hyposmia, which is similar to prior report^29^ and, as expected, lower than the rate (60-90%) of objective hyposmia observed in iRBD or PD patients. Subjective hyposmia alone has limited sensitivity in iRBD, however combined with actigraphy, specificity reached 100% in our sample. Although objective measurement of olfaction by validated smell identification kits is more sensitive, subjective assessment of hyposmia could significantly reduce cost of material, shipment, as well as burden on participants and providers in the setting of a large population screen.

Altogether, our findings suggest that a comprehensive iRBD/prodromal synucleinopathy questionnaire could improve current clinical diagnostic procedure. A two-step approach using questionnaire first, followed by ambulatory actigraphy in those who screen positive, could select individuals at highest risk of iRBD, and thereby result in higher positive predictive value (100% in this small sample of 84 subjects), before confirmatory vPSG. Although performance may be lower in population groups with a high prevalence of sleep disorders, it is expected to perform as well or better in non-clinical populations expected to have less sleep comorbidities than patients seen at a sleep clinic. Future studies are needed to test this ambulatory screening procedure in large sample of the general population.

Limitations in this study include its small sample size, which could reduce the robustness of our findings. Clinic CL had a relatively low representation of “pseudo-RBD” secondary to OSA, and post-traumatic stress disorder (PTSD), which can also mimic RBD. The study was not powered to thoroughly evaluate performance across sleep disorders.

The RBD set, in turn, was well-characterized and consistent with the classic description of “idiopathic” RBD, associated in 79 % of cases with at least one other disease prodrome, and vPSG finding of prominent upper extremity and chin EMG augmentation. One misclassified patient was a female with no other prodromes of synucleinopathy and near-complete absence of arm EMG elevation on vPSG, raising the possibility that upper extremity RSWA is necessary for actigraphy detection. The second was a man with borderline RSWA. Studies are needed to explore the relation between severity and topographic distribution of RSWA, and performance of wrist actigraphy.

Studies should also be conducted to test the utility of the actigraphy model for detection of RBD associated with overt synucleinopathy, or other RBD phenotypes such as RBD associated with narcolepsy and post-traumatic stress disorder.

Our detection paradigm has the potential to be used in a staged approach, using questionnaires first, followed by actigraphy in positive screens, to select individuals with high pretest probability before confirmatory vPSG. This algorithm could be implemented fully remotely in outpatient clinics. The study also lays the ground for future population studies aiming at developing precise and cost-effective screening strategies for iRBD in the general population.

## Data Availability

All data produced in the present study are available upon reasonable request to the authors-

