## Supplementary Information for "Ambulatory detection of isolated REM sleep behavior disorder combining actigraphy and questionnaire" (from another version of this preprint: version 2, 2022-04-27)

**1 eMethods**

**1.1 3-item Questionnaire on Nonmotor Symptoms of Synucleinopathy**

1) *Hyposmia*: “*Have you been told, or have you noticed that your sense of smell or taste is reduced compared to others or to what it used to be?”*
2) Constipation*: “Do you experience constipation (difficulty having a bowel movement every day or having to strain hard to pass stools) OR need to use laxatives or other treatments to improve your bowel transit?”*
3) Orthostasis*: “In the past month, when standing up have you had the feeling of either becoming lightheaded, or no longer being able to see properly, or no longer being able to think clearly?”*

**1.2 Activity Count**

The accelerometer data was resampled to exactly 25 Hz using cubic spline interpolation^29^. The accelerometer data was calibrated such that the vector magnitude is equal to the gravitational acceleration (1 g) in periods of inactivity. This calibration was implemented with an iterative closest-point fitting process that previously have been shown to reduce error^30^. Activity counts were calculated as the acceleration magnitude

$$Activity Count_{i}=\left| \sqrt{x_{i}^{2}+y_{i}^{2}+z_{i}^{2}}-1 \right|,$$

Where $x_{i}$, $y_{i}$, and $z_{i}$ are acceleration in the three axes. The activity counts were then averaged in 1-second, 15-second or 30-second bins. To avoid baseline drift in the activity counts, they were forward-backward filtered using a high-pass infinite impulse response (IIR) Butterworth filter with a stopband attenuation of 80 dB at 0.1 Hz and passband ripple of 1 dB starting at 0.5 Hz. Finally, activity counts less than 0.1 were rounded to zero.

**1.3 Ensemble of Decision Trees**

The ensemble of decision trees consisted of 400 boosted trees that were optimized with LogitBoost^31^ using a learning rate of 0.1. The decision trees used all features, a maximum of 10 splits, a minimum of 1 leaf, and used the Gini’s diversity index (gdi) as a split-criterion.

**1.4 Evaluation of Feature Importance**

The feature importance (FI) in the decision tree ensembles were estimated by considering changes in node risks for each feature. The node risk $R_{i}$ is defined as

$$R_{i}=P_{i}\cdot gdi_{i},$$

where $P_{i}$ is the probability of reaching node $i$ and $gdi_{i}$ is the gdi at node $i$. The change in node risk at node $i$ is

$$R_{\Delta i}=R_{i}-R_{C1}-R_{C2},$$

where the subscript $C1$ and $C2$ denotes the child nodes. Finally, the node risk changes are averaged across features. The FI of feature k is defined as

$$FI_{k}=\frac{\sum_{i\epsilon N_{k}} R_{\Delta i}}{\left| N_{k} \right|},$$

where $N_{k}$ is the set of nodes using feature $k$.

**2 eFigures**

**eFig. 1: Performance for actigraphy-based classifier as a function of number of nights recorded.**

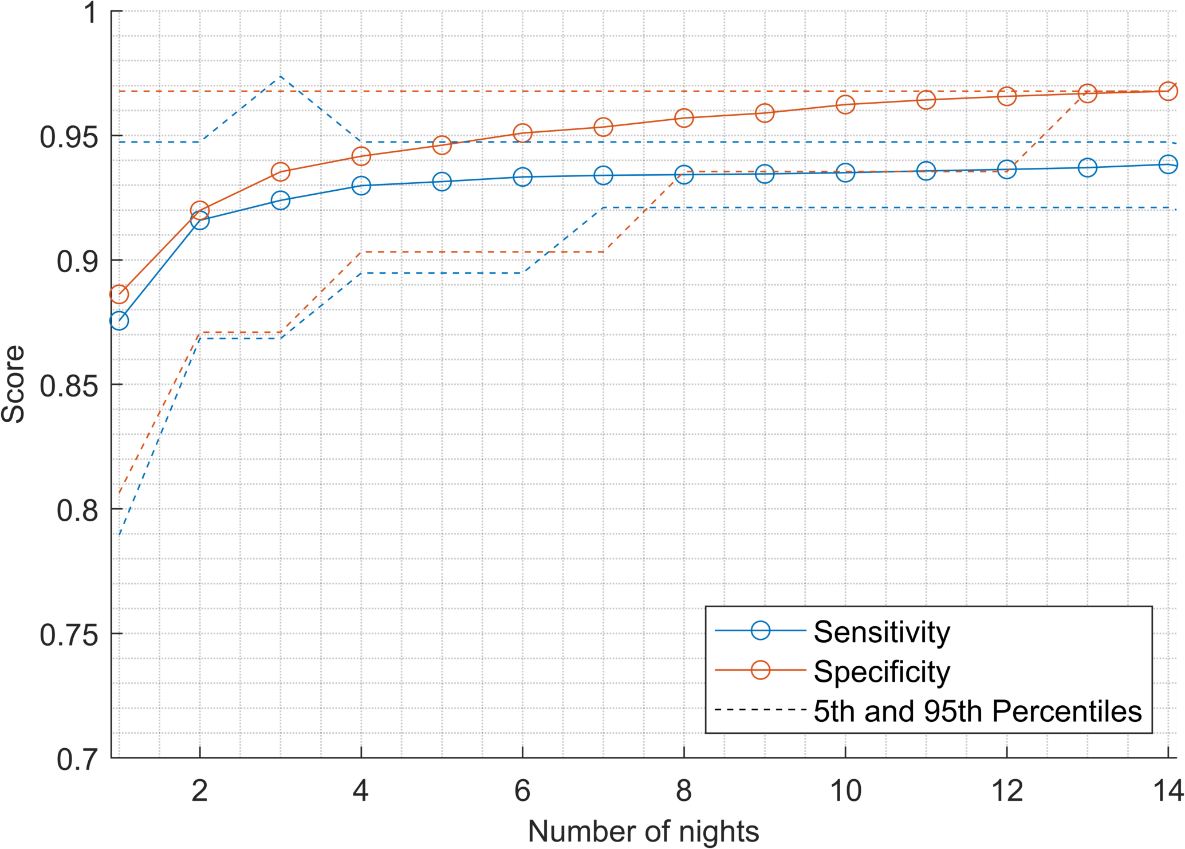

The performance seems to saturate after recording 7 to 10 nights. The performance was evaluated in a subset with at least 14 nights, which includes 18 controls and 32 iRBD participants. The solid lines indicate the mean while the dashed lines indicate 5^th^ and 95^th^ percentiles estimated through bootstrapping.

**eFig. 2: Feature importance of the actigraphy-based classifier.**

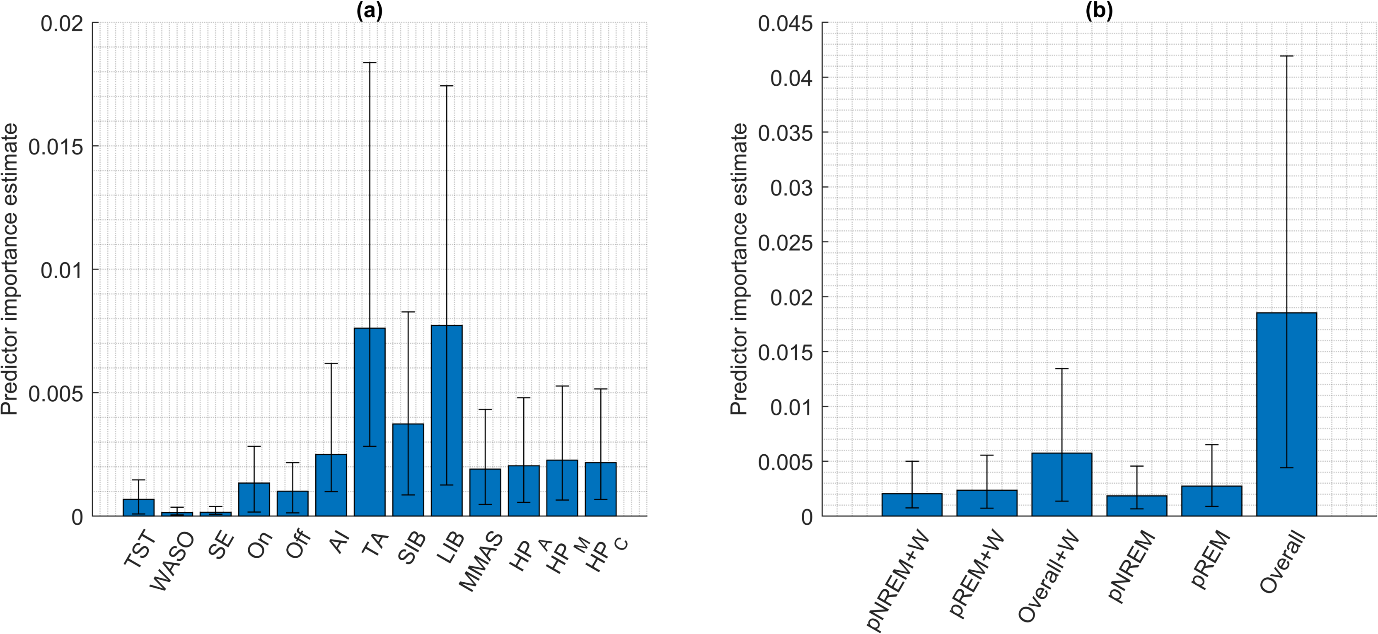

Feature importance was estimated in the boosted decision trees that used actigraphy features by weighing the importance of each split. These were grouped and added by **(a)** feature type and **(b)** feature mask. TST: total sleep time; WASO: wake after sleep onset; SE: sleep efficiency; On: sleep onset; Off: sleep offset; AI: activity index; TA: twitch activity; SIB: short immobile bouts; LIB: long immobile bouts; MMAS: mean motor activity score; HP_A_: Hjorth activity parameter; HP_M_: Hjorth mobility parameter; HP_C_: Hjorth complexity parameter.

**eFig. 3: Feature importance of the questionnaire-based classifier.**

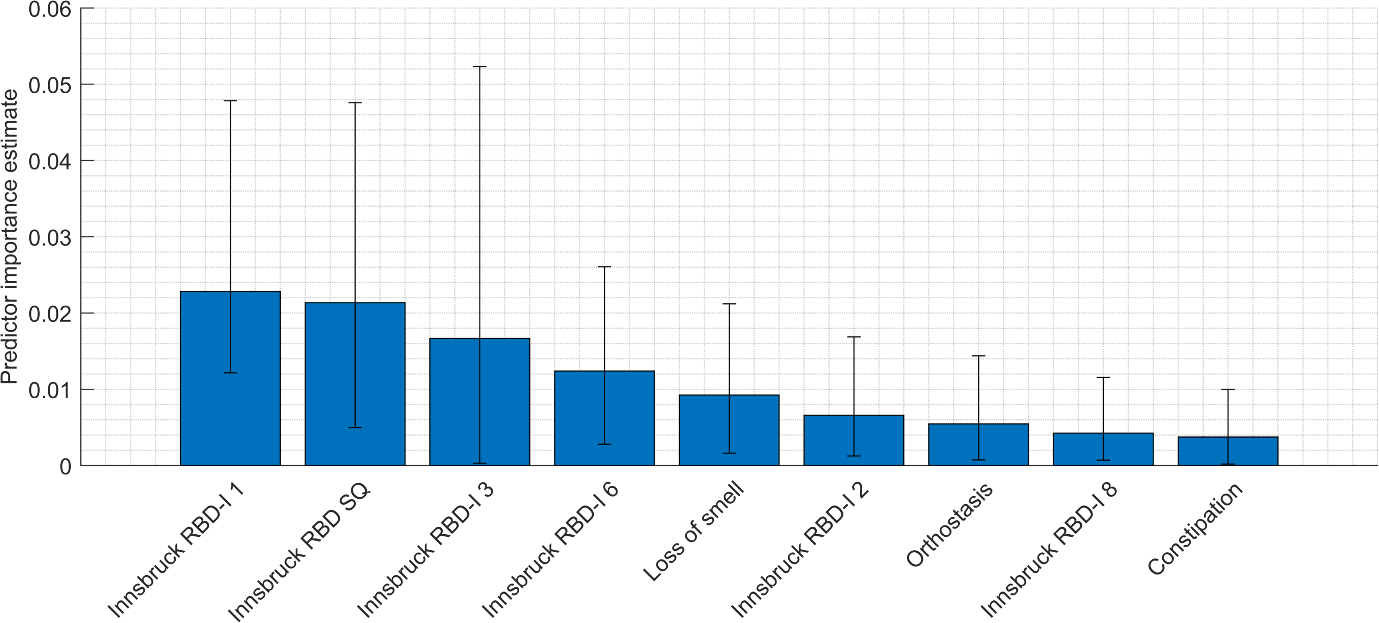

Feature importance was estimated in the boosted decision trees that used actigraphy features by weighing the importance of each split.

**3 eTables**

**eTable 1: Most discriminative actigraphy features of each type.**

|  |  | Control | Case | p-val |
| --- | --- | --- | --- | --- |
|  | n | 42 | 41 |  |
| AI (x = 60) | μ ± σ | 0.196 ± 0.057 | 0.379 ± 0.122 | **1.53×10^-11^** |
| TA (x = 0.5) | μ ± σ | 7.25 ± 2.52 | 18.6 ± 8.95 | **5.56×10^-12^** |
| SIB (x = 5) | μ ± σ | 0.345 ± 0.044 | 0.455 ± 0.073 | **3.66×10^-11^** |
| LIB (x = 60) | μ ± σ | 6.32 ± 1.50 | 10.3 ± 1.96 | **1.62×10^-12^** |
| MMAS_pREM_ | μ ± σ | 0.021 ± 0.009 | 0.031 ± 0.015 | **9.87×10^-5^** |
| HP_A; pREM_ | μ ± σ | 0.064 ± 0.038 | 0.091 ± 0.062 | **0.0141** |
| HP_M; pREM_ | μ ± σ | 0.954 ± 0.052 | 0.992 ± 0.045 | **0.00137** |
| HP_C_ | μ ± σ | 1.69 ± 0.066 | 1.64 ± 0.048 | **0.000715** |

Statistics are computed using Mann-Whitney tests for all features. In the feature names, x denotes a parameter of the feature. Here, we show the distribution of features with the best parameter and feature mask determined by lowest *p*-value. *P*-values < 0.05 are considered significant and marked in bold. The subscripts NREM, REM, and w denotes the feature masks of the feature. AI: activity index; TA: twitch activity; SIB: short immobile bouts; LIB: long immobile bouts; MMAS: mean motor activity score; HP_A_: Hjorth activity parameter; HP_M_: Hjorth mobility parameter; HP_C_: Hjorth complexity parameter.

**eTable 2: Questionnaires for REM sleep behavior disorder and other synucleinopathy prodromes.**

|  |  | CL | iRBD | *p*-value | Acc |
| --- | --- | --- | --- | --- | --- |
|  | n | 42 | 41 |  |  |
| Innsbruck RBD SQ | n, (%) | 3, (7.14 %) | 35, (87.5 %)  (n = 40) | **3.01×10^-13^** | 90.2 % |
| Innsbruck RBD-I | μ ± σ | 0.08 ± 0.21 | 0.80 ± 0.27 | **1.15×10^-13^** | 90.4 % |
| - Item 1 | n, (%) | 3, (7.14 %) | 35, (92.1 %)  (n = 38) | **2.98×10^-14^** | 92.5 % |
| - Item 2 | n, (%) | 4, (9.52 %) | 25, (67.6 %)  (n = 37) | **9.25×10^-8^** | 79.8 % |
| - Item 3 | n, (%) | 2, (4.88 %)  (n = 41) | 33, (91.7 %)  (n = 36) | **2.33×10^-14^** | 93.5 % |
| - Item 6 | n, (%) | 4, (9.52 %) | 28, (70.0 %)  (n = 40) | **2×10^-8^** | 80.5 % |
| - Item 8 | n, (%) | 3, (7.50 %)  (n = 40) | 21, (80.8 %)  (n = 26) | **1.48×10^-9^** | 87.9 % |
| 3Q | μ ± σ | 0.10 ± 0.23 | 0.51 ± 0.34 | **1.21×10^-7^** | 78.3 % |
| - Hyposmia | n, (%) | 3, (7.14 %) | 22, (57.9 %)  (n = 38) | **1.01×10^-6^** | 76.3 % |
| - Constipation | n, (%) | 5, (11.9 %) | 25, (61.0 %) | **3.28×10^-6^** | 74.7 % |
| - Orthostasis | n, (%) | 5, (11.9 %) | 14, (35.0 %)  (n = 40) | **0.0132** | 62.2 % |

Statistics are computed using Mann-Whitney test for Innsbruck RBD-I and chi-squared tests for other variables. If participants answered, “don’t know”, the data is regarded as missing, hence, the sample sizes are varying. The accuracy (Acc) was calculated using the cut-off of 0.25 for the RBD-I as it was the optimal threshold in the original publication^20^. The Acc for the 3Q was calculated using an optimal cut-off of 0.33 (at least one positive question). *P*-values < 0.05 are considered significant and marked in bold. RBD SQ: RBD summary question; RBD-I: RBD inventory.

**eTable 3: Performance in classification of isolated REM sleep behavior disorder using optimized thresholds.**

|  | Sensitivity  (95 % CI) | Specificity  (95 % CI) | Accuracy  (95 % CI) | Precision  (95 % CI) |
| --- | --- | --- | --- | --- |
| Actigraphy |  |  |  |  |
| Multi-feature |  |  |  |  |
| Per night | 90.0 (81.9 - 95.7) | 85.2 (76.1 - 92.3) | 88.0 (79.0 - 94.1) | 89.6 (80.4 - 94.9) |
| Per subject | 95.1 (88.1 - 98.7) | 95.2 (88.1 - 98.7) | 95.2 (88.1 - 98.7) | 95.1 (88.1 - 98.7) |
| Single-feature (SIB) | | | | |
| Per night | 79.6 (69.2 - 87.6) | 76.7 (66.6 - 85.6) | 78.4 (67.9 - 86.6) | 82.9 (73.3 - 90.5) |
| Per subject | 85.4 (76.1 - 92.3) | 88.1 (79.0 - 94.1) | 86.7 (77.5 - 93.2) | 87.5 (79.0 - 94.1) |
| Questionnaire |  |  |  |  |
| RBD-I | 90.2 (81.9 - 95.7) | 92.9 (84.9 - 97.3) | 91.6 (83.4 - 96.5) | 92.5 (84.9 - 97.3) |
| RBD-I + 3Q | 97.6 (91.6 - 99.7) | 85.7 (76.1 - 92.3) | 91.6 (83.4 - 96.5) | 87.0 (77.5 - 93.2) |
| 3Q | 75.6 (65.3 - 84.6) | 88.1 (79.0 - 94.1) | 81.9 (72.0 - 89.5) | 86.1 (76.1 - 92.3) |
| Actigraphy + Questionnaire | 92.7 (84.9 - 97.3) | 100.0 (95.7 - 100.0) | 96.4 (89.8 - 99.2) | 100.0 (95.7 - 100.0) |

The performance is reported for classification based on actigraphy, questionnaires, and the combination of these. The combination classifies a sample as belonging to the iRBD or CL group if actigraphy and questionnaire predictions are concordant. The 95 % CI were estimated using the Clopper-Pearson method. CI: confidence interval.

**eTable 4: Performance in classification of isolated REM sleep behavior disorder using 15-second activity counts.**

|  | Sensitivity  (95 % CI) | Specificity  (95 % CI) | Accuracy  (95 % CI) | Precision  (95 % CI) |
| --- | --- | --- | --- | --- |
| Actigraphy |  |  |  |  |
| Multi-feature |  |  |  |  |
| Per night | 89.1 (80.4 - 94.9) | 85.3 (76.1 - 92.3) | 87.5 (79.0 - 94.1) | 89.6 (80.4 - 94.9) |
| Per subject | 95.1 (88.1 - 98.7) | 88.1 (79.0 - 94.1) | 91.6 (83.4 - 96.5) | 88.6 (80.4 - 94.9) |
| Single-feature (SIB) | | | | |
| Per night | 76.5 (65.3 - 84.6) | 71.1 (60.1 - 80.5) | 74.2 (64.0 - 83.6) | 79.0 (69.2 - 87.6) |
| Per subject | 90.2 (81.9 - 95.7) | 73.8 (62.7 - 82.6) | 81.9 (72.0 - 89.5) | 77.1 (66.6 - 85.6) |
| Actigraphy + Questionnaire | 87.8 (79.0 - 94.1) | 97.6 (91.6 - 99.7) | 92.8 (84.9 - 97.3) | 97.3 (91.6 - 99.7) |

The performance is reported for classification based on actigraphy, questionnaires, and the combination of these. The combination classifies a sample as belonging to the iRBD or CL group if actigraphy and questionnaire predictions are concordant. The 95 % CI were estimated using the Clopper-Pearson method.

**eTable 5: Performance in classification of isolated REM sleep behavior disorder using 30-second activity counts.**

|  | Sensitivity  (95 % CI) | Specificity  (95 % CI) | Accuracy  (95 % CI) | Precision  (95 % CI) |
| --- | --- | --- | --- | --- |
| Actigraphy |  |  |  |  |
| Multi-feature |  |  |  |  |
| Per night | 89.8 (81.9 - 95.7) | 85.8 (76.1 - 92.3) | 88.1 (79.0 - 94.1) | 89.9 (81.9 - 95.7) |
| Per subject | 95.1 (88.1 - 98.7) | 88.1 (79.0 - 94.1) | 91.6 (83.4 - 96.5) | 88.6 (80.4 - 94.9) |
| Single-feature (SIB) | | | | |
| Per night | 83.6 (73.3 - 90.5) | 81.3 (70.6 - 88.6) | 82.7 (73.3 - 90.5) | 86.4 (77.5 - 93.2) |
| Per subject | 95.1 (88.1 - 98.7) | 78.6 (67.9 - 86.6) | 86.7 (77.5 - 93.2) | 81.3 (70.6 - 88.6) |
| Actigraphy + Questionnaire | 87.8 (79.0 - 94.1) | 100.0 (95.7 - 100.0) | 94.0 (86.5 - 98.0) | 100.0 (95.7 - 100.0) |

The performance is reported for classification based on actigraphy, questionnaires, and the combination of these. The combination classifies a sample as belonging to the iRBD or CL group if actigraphy and questionnaire predictions are concordant. The 95 % CI were estimated using the Clopper-Pearson method.

**eTable 6: Correlation between actigraphy iRBD score and demographics and clinical variables.**

|  | iRBD |  | Control |  |
| --- | --- | --- | --- | --- |
|  | *r* | *p*-val | *r* | *p*-val |
| Age | 0.05 | 0.75 | 0.17 | 0.28 |
| BMI | 0.22 | 0.16 | 0.03 | 0.83 |
| Sex (% male) | 0.24 | 0.13 | 0.01 | 0.97 |
| Antidepressants | 0.20 | 0.2 | -0.10 | 0.53 |
| Melatonin | -0.09 | 0.58 | - | - |
| Clonazepam | 0.20 | 0.22 | -0.02 | 0.88 |
| Rivastigmine | -0.13 | 0.43 | - | - |
| Pramipexole | 0.22 | 0.16 | - | - |
| PLMi  (Control: *n* = 19) | 0.05 | 0.75 | 0.02 | 0.93 |
| OSA on PAP therapy | **-0.42** | **0.022** | 0.30 | 0.24 |
| AHI in untreated OSA  (Controls: *n* = 17, iRBD: *n* = 30) | 0.27 | 0.086 | 0.14 | 0.39 |
| RLS/PLM | 0.01 | 0.97 | -0.13 | 0.41 |
| NREM parasomnia | 0.05 | 0.75 | -0.03 | 0.84 |
| Hypersomnia | 0.22 | 0.17 | **0.36** | **0.019** |
| Insomnia | -0.11 | 0.49 | **0.35** | **0.018** |
| Parasomnia episodes per night | 0.18 | 0.26 | 0.20 | 0.2 |
| RSWAi  (iRBD: *n* = 35) | -0.02 | 0.91 | - | - |

The correlation was computed as Pearson’s correlation coefficient. *P*-values < 0.05 are considered significant and marked in bold.
